# The Digital Health Revolution: Exploring the Impact of Online Cancer Information on Self-Reported Preventive Behaviors

**DOI:** 10.1101/2024.05.20.24307517

**Authors:** Bryan Yavari, Nilofar Kolbehdari, Lindsay Gann, Mercedes Portillo, Alexandria Rumschlag, Melanie Aldridge, Walker Mellon, Gissel Marquez Alcaraz, Harley Richker, Mané Sarkissian, Zachary T. Compton, Athena Aktipis, Carlo Maley, Cristina Baciu

## Abstract

Cancer, one of the leading causes of death worldwide, is a disease characterized by uncontrolled cell growth within the body. While there have been many improvements in the treatment of cancer clinically, there is now an urgent need to improve cancer-related communication. This study explores the impact of online health information, specifically cancer-related information and prevention, among members of the general public. Through a randomized survey, we examined what information leads people to take action to minimize their cancer risk and communicate with their providers. Through evaluation of the various modes of communication, we were able to provide insight into which are more effective and better received by members of the general public. Through this, ways of bettering these avenues of communication and strengthening the bond between them will be highlighted and more easily elaborated on by future studies. The results of our study indicated that 60% of participants asserted that they are motivated by online preventive information to take steps to limit their cancer risk, while only roughly 44% of participants overall agreed that their doctor has communicated with them about when proper cancer screenings should be scheduled for the future. Although patients may be turning to the Internet now more than ever due to various reasons, when comparing self-reported rates of comprehension among the study participants, 35% agreed that the cancer-related information they can access online is confusing, while fewer than 22% of participants agreed that the cancer-related information they receive directly from their doctor is confusing. This is indicative of the limitations the Internet may have when undertaking the role of being a medical resource, especially when acting as a replacement for in-person medical appointments where patients can communicate directly with their physicians. Ultimately, these results provide a unique perspective into how people receive, evaluate, and implement cancer-preventive steps and general health-related information in a post-COVID-19 world, where the Internet is now strongly embedded in healthcare.

## Introduction

This study aims to examine the impact of online cancer information on the overall patient experience and perception of the general public. Factors affecting this impact, such as accessibility to the Internet and how the public utilizes this information, are investigated throughout this study. Although there have been previous studies on this topic, there has been limited research on cancer communication, specifically online communication, after COVID-19. The pandemic has shifted much of this communication to virtual sources, highlighting the importance of this topic.

Cancer, one of the leading causes of death in the United States (Ahmad & Anderson, 2021), is an important component of public health and is a topic of longstanding informational media campaigns (Klein et al., 2014). Over the last decade, cancer- and health-related information has surfaced in ways that make it instantly accessible, namely through the unrestricted Internet access available to the greater population (Fareed et al., 2021). Not all publicly available cancer prevention information, however, is reliably sourced, updated, or accurate, resulting in poor comprehension and applicability to the viewer.

Recent times have seen an increase in the use of the Internet for the identification of illness-related symptoms and consequent diagnoses, measuring the degree of risk factors for various health-related behaviors, and broadcasting preventive information. In light of this, it is important for the individuals accessing this information to be able to assess the validity of what they are viewing. The ability to differentiate between accurate and inaccurate online health-related information is a concept known as health literacy, something that has been found to be low among the general public (Diviani et al., 2015). Easy and immediate accessibility is a key factor that plays a role in the rising rates of Internet usage for cancer and overall health-related information. With varying and limited capabilities for evaluating the credibility of online resources among members of the public, many fall victim to the rise in misinformation media campaigns (Mantwill et al., 2015).

Trust and confidence, on the other hand, have additionally been correlated as two of the main factors predicting the type of resources an individual may gravitate toward when seeking health-related information (Jacobs et al., 2017). A patient’s trust in their physician is crucial for desirable treatment and prevention outcomes such as satisfaction and adherence to treatment plans. In oncology, trust is possibly even more essential due to the life-threatening nature of cancer (Hillen et al., 2011). When this sense of trust between patient and provider is lacking, people often turn to what feels like their only other option, the Internet (J. K. Davis, 2018).

Welcome or not, this new age of technology is making waves in the world of medicine. Providing critical information to members of the public with limited access to traditional medical resources, the various benefits of readily available telehealth appointments, and the widespread reach of prevention and risk-factor information are only a few of the numerous positives the Internet has brought to the medical field over the last several years (Ahmadi et al., 2019). With it, however, comes the spread of misinformation, the exploitation of the lack of health literacy among the general public, and an increased sense of mistrust and doubt between patients and providers (Steinke & Hövelmann, 2020).

### The Rapid Rise in Online Cancer-Related (Mis)Information

By completing a comprehensive literature review before phasing into the data collection phase of the study, a basis of focus was established, bringing to light the key aspects of the spread of online cancer-related information and the gaps that currently exist in the literature. In a cross-sectional survey completed by 635 participants in 2013, the Internet was reported to be the number one source of health-related information among 82.7% of respondents (Van de Belt et al., 2013). The Internet continues to be the number one most used resource for health information (Pan et al., 2021). Over the last few years, especially after the events surrounding the COVID-19 pandemic, society has continued to delve deeper into the world of technology, further maximizing the role of the Internet on health-related behaviors. With little doubt about the growing impact of the Internet, the scientific community has taken notice of this new key player within the world of medicine. However, the large amount of readily available yet inaccurate information leads to a higher potential for spreading misinformation (Swire-Thompson & Lazer, 2020).

A recent meta-analysis suggests that the number of research studies that investigated health-related misinformation increased over the years, from 7 in 2012 to 41 just six years later in 2018, with a sharp rise in misinformation in 2017 (Y. Wang et al., 2019). Among the increasing number of studies conducted, however, there is a general consensus that the spread of health-related misinformation on social media networks tends to be more prevalent than scientifically accurate knowledge, inducing medical-related fear, anxiety, and mistrust among the general public (K. Kim et al., 2023).

### Misinformation Presents Challenges in Following Accurate Health Guidelines

In what is being called “the era of fake news,” misinformation has been found to spread expeditiously through the general public via the internet. This brings to light a major concern regarding whether any online cancer information is accurate, up-to-date, and trustworthy. Taking it one step further, in a recently conducted study, videos and social media content containing cancer-health information constitute 76.9% of the misleading information and can lead to incorrect healthcare proceedings or impede treatment plans (Teplinsky et al., 2022). A lack of health literacy renders members of the general public even more susceptible to this spread of misinformation, leading to potentially detrimental circumstances in their health (K. Kim et al., 2023).

Additionally, individuals lacking general literacy consequently have difficulties understanding common medical language, such as medical terminology, testing results, or screening recommendation processes (H. Kim & Xie, 2017). Left to their own accord with no guidance or explanation from a healthcare provider, many in the population then turn to the Internet where they are unable to properly decipher what they are reading or the results they may be receiving through an online portal.

### Americans are Behind in Health Literacy, Causing a Delay in Action

Despite its pitfalls, the Internet has now been recognized worldwide as an important conduit for improving overall health literacy among the public. Health literacy is defined as “the degree to which individuals have the capacity to obtain, process, and understand basic health information and services needed to make appropriate health decisions” (Rudd et al., 2023). Unfortunately, nearly half of American adults have only basic health literacy, and similar levels have been found in Europe and other developed countries (S. Jiang & Beaudoin, 2016). In a clinical setting, such individuals with low health literacy rates are at a significant disadvantage in their ability to process, understand, and act on the cancer information and services needed to make appropriate healthcare decisions. Because of this, health literacy is increasingly recognized as a critical factor affecting communication across the continuum of cancer care (Ryman et al., 2024).

By cultivating critical thinking and taking steps to improve overall health literacy among the public, individuals will be better equipped to communicate with their healthcare providers, understand the health-related information they see online, and practice an overall healthy lifestyle. The Internet and social media have the potential to be tools that can be used to empower patients, allowing them to feel more in control even when facing a serious health concern such as cancer.

### Maximizing the Spread of Prevention Strategies is Crucial to Early Detection

Although steps are being taken every day to find the “cure” to cancer, prevention remains the most cost-effective and efficient strategy to fight cancer. For decades, public health efforts have used mass media to provide information to the public and raise awareness about lifestyle cancer-related behaviors in hopes of encouraging them to make more improved lifestyle choices. More often than not, however, health promotion advertisements include imagery and anecdotes that induce high levels of negative emotion in viewers. Although the goal is to decrease rates of health-compromising behaviors such as smoking, the resulting negative emotions experienced by some viewers can induce overall resistance to the message of the advertisement rather than compliance (Di Giuseppe et al., 2019).

On the other hand, in an analytical study conducted in 2019, cancer prevention information that contained both high levels of threat and efficacy gained the largest number of readings and likes on social media platforms (X. Wang et al., 2019). This discrepancy in the literature further highlights the need for additional research on the most beneficial way to advocate for increased cancer-related information and prevention behaviors among the general public when utilizing mass media as a tool of communication.

### The Importance of Cultivating a Healthy Relationship Between Patient & Provider

Although the Internet may have a wider reach when it comes to advocating prevention strategies and cancer-related information, many still believe that the best recommendations come directly from their healthcare providers. In a previously conducted study of 294 participants, 86% reported that they would prefer to receive any health-related information from a healthcare professional (Wainstein et al., 2006). However, in a study of 704 breast cancer patients, only 65% reported that they always trust their provider, with similar levels of trust across patients with other conditions (Kaiser et al., 2011).

In a healthcare system where it is oftentimes difficult to get an appointment with a provider and even more challenging to pay for the resulting medical expenses, establishing a sense of trust and providing an environment where patients can have confidence in the information being provided to them should be a priority for any healthcare provider and establishment. Here, we assessed the current opinions on online cancer communication and clinical interactions.

## Methodology

### Quantitative and Qualitative Results Reveal Various Opinions

This comprehensive quantitative and qualitative research study began through an extensive literature review of peer-reviewed entries that contain validated survey questions. We analyzed these questions in conjunction with previously published scientific papers to explore health literacy, online health information, and existing connections between online information and cancer-preventive behavior. The survey questions in this study were created to fill existing gaps in this knowledge that were created by the COVID-19 pandemic. These gaps include addressing the increased use of telehealth and online searches relating to cancer-preventive information.

The survey for this study consisted of 10 Likert Scale questions (1 to 5) and five open-ended questions to detail the specific and broad influence of this information on preventive actions. The questions were designed with objectivity and simplicity in mind to limit all possible biases. Many of the questions were validated questions from previously delivered surveys to obtain new information in today’s new online climate.

### Variation in Surveyed Population Provides Unique Data

Anyone in the general public over the age of 18 was eligible to be part of this study. The survey was distributed online through Prolific, a high-quality online research platform that screens participants to connect them with different surveys. We received a large diversity of respondents with regard to age, income, location, marital status, and education level.

We thoroughly analyzed this survey’s data to assess the mean, standard deviation, and correlational analyses. The Likert Scale questions provided a quantifiable comparison among the general public and a comparison to varying demographic factors. The open-ended questions were analyzed in depth to assess the recurrent answers and identify themes. These answers were then grouped and analyzed to determine respondents’ behaviors and practices.

### COVID-19 Has Changed People’s Opinions and Actions

Our mixed-methods approach created a multifaceted examination of the fundamental impact of online cancer information on preventive health behaviors. These insightful responses and analyses deepened our understanding of how online cancer information drives current preventive behaviors, especially after the COVID-19 pandemic.

## Results

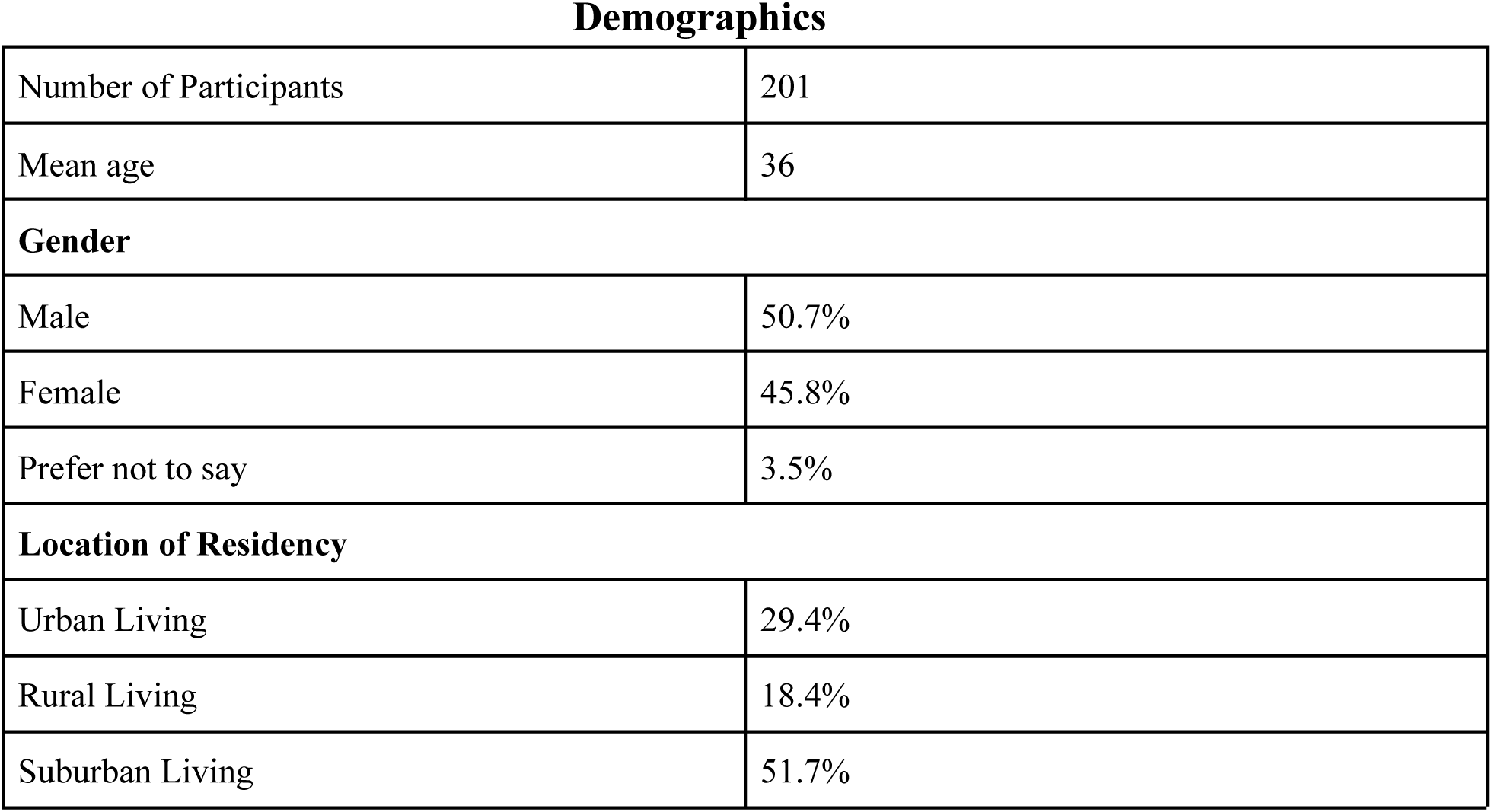
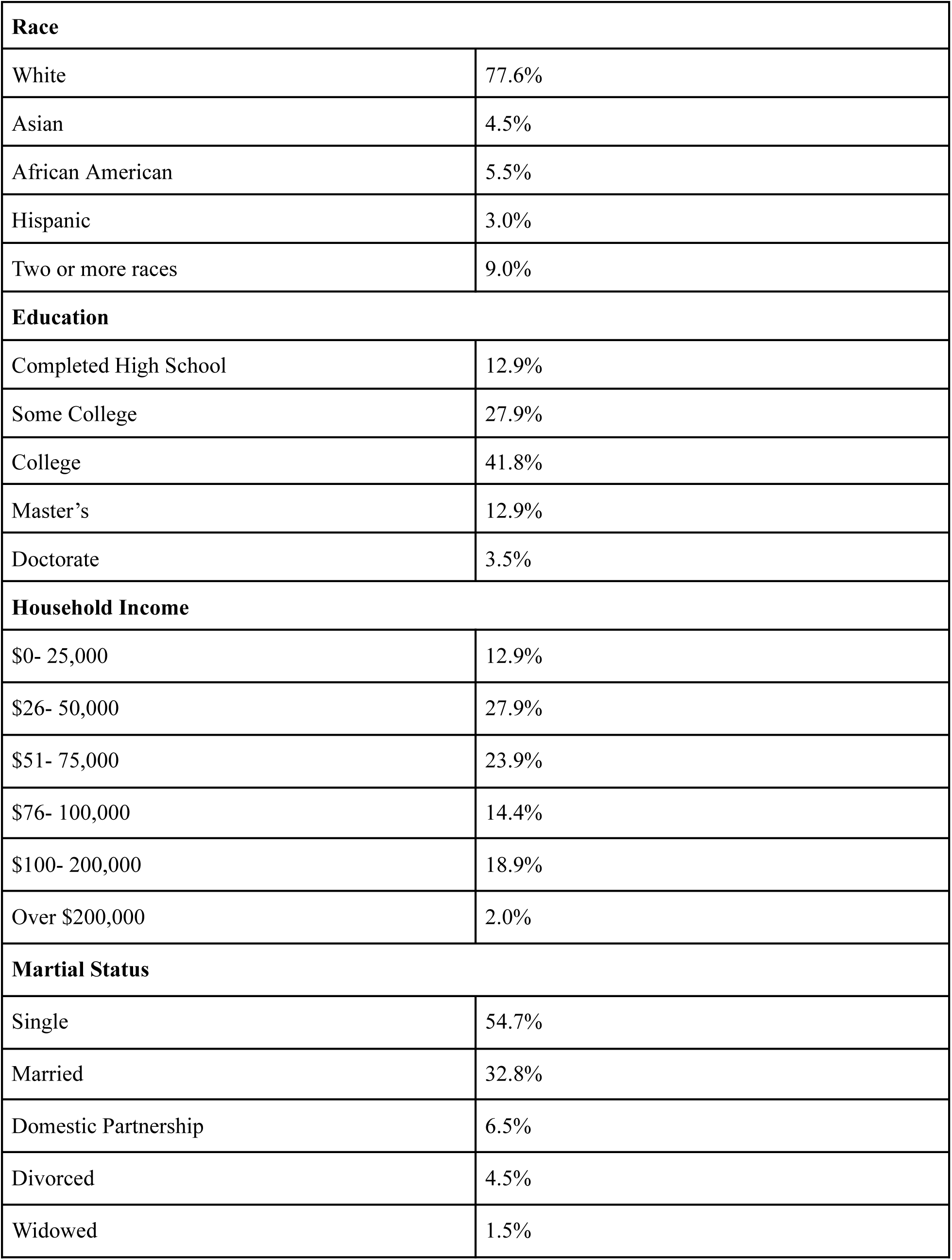
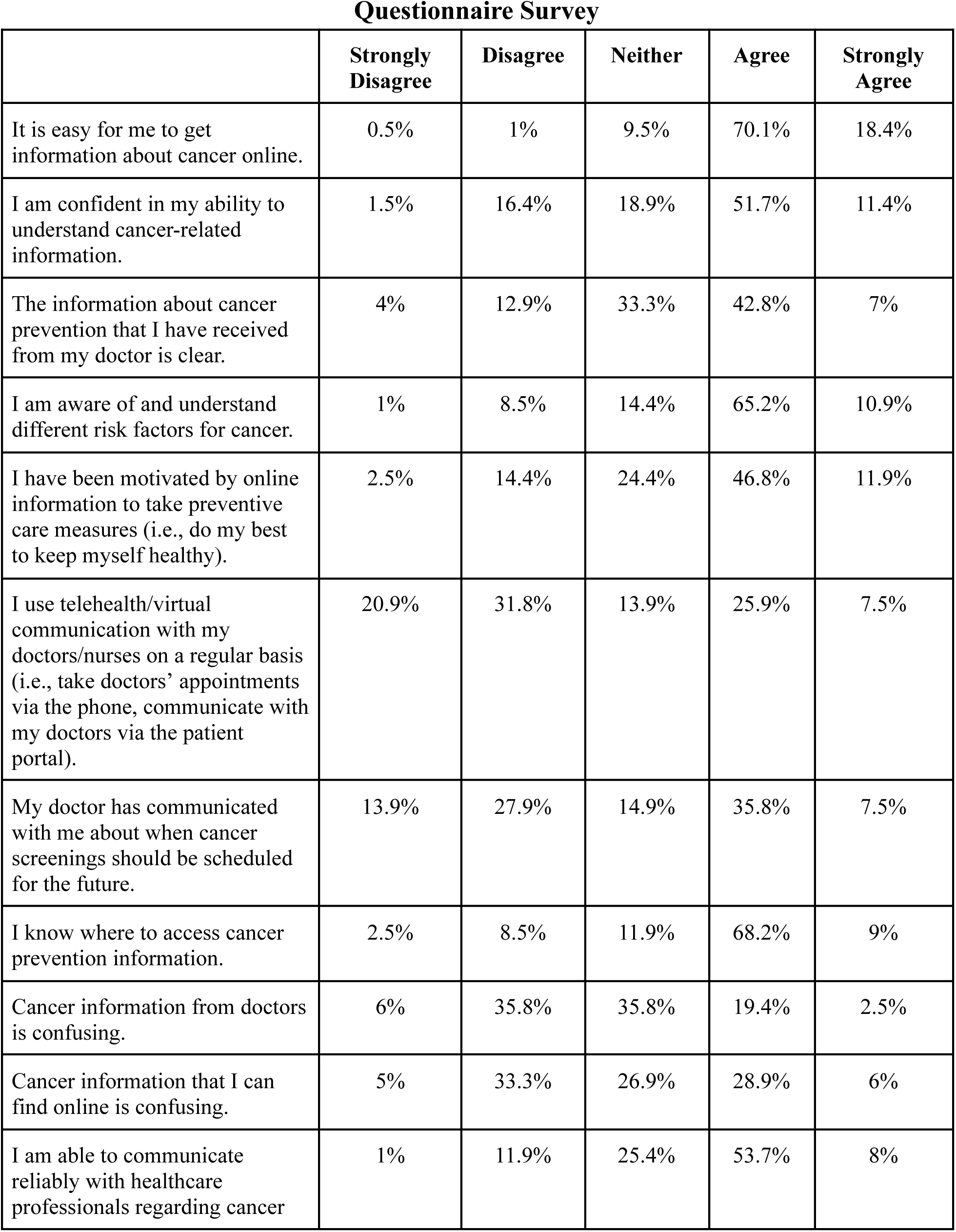
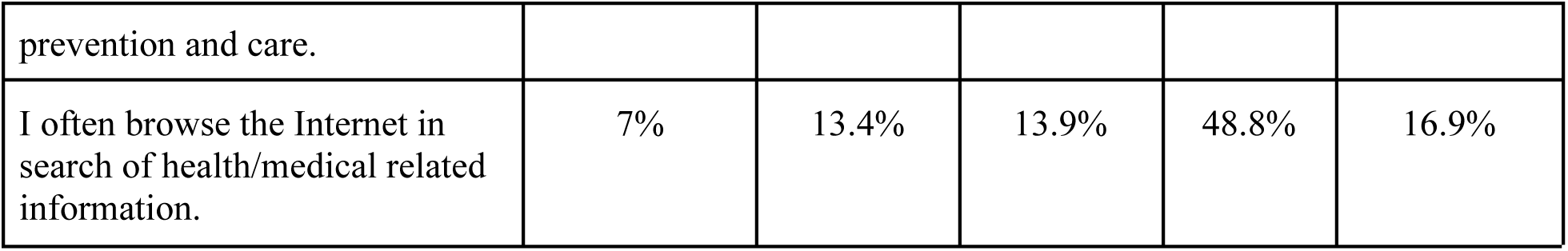

### Communication and Comprehension of Cancer is Crucial for Patient-Provider Relationships

Conducting the survey with the general public helped our team better understand the public’s attitudes toward cancer-preventive information. Our results indicated that 70 percent of respondents agree that it is easy to get cancer information online, with 18 percent strongly agreeing. Additionally, just 43 percent of respondents agree, and 7 percent strongly agree that the information about cancer prevention they receive from their doctor is clear.

On the other hand, approximately 21 percent of participants agreed and strongly agreed that the cancer information from their doctor is confusing, while approximately 35 percent of participants agreed overall that the cancer information they access online is confusing. Although cancer-related information is easier to access on the internet than in-person appointments, our study has found that respondents often find it more difficult to comprehend.

Our team was further interested in assessing if the general population feels that cancer is talked about enough. We asked, “Is cancer talked about too much or too little in public?” Roughly 60 percent of respondents believed that cancer is talked about too little in public, while about 10 percent stated it was talked about just right, and the other 30 percent said cancer is talked about too much.

To gain insight into communication regarding regular screenings and checkups, our survey also asked: “My doctor has communicated with me about when cancer screenings should be scheduled for the future.” The responses to this question revealed that 14 percent of respondents strongly disagreed, 28 percent disagreed, and 15 percent were neutral. With roughly 57 percent of respondents ultimately not agreeing that their doctor communicates with them about when cancer screenings should be scheduled, it reveals why many people turn to online resources to find cancer information.

### Online Cancer Information May Lead to Improved Preventive Care

Our survey asked respondents: “I have been motivated by online information to take preventive care measures (i.e., do my best to keep myself healthy).” The results indicated that 59% of respondents agreed or strongly agreed, 24% were neutral, and only 16% disagreed or strongly disagreed. Our findings suggest that people are motivated and able to search for cancer information online and implement preventive measures. Another question led to 66% of the respondents agreeing that they know where to access cancer prevention information.

Health-related information provided to patients over the Internet provides cancer patients with resources for their care and treatment, improving patient care. This study highlighted the rapid projection of cancer prevention information obtained through the internet and more recently, social media platforms. When our participants were asked if they often browse the Internet for medical-related information, approximately 49% agreed, and 17% strongly agreed. With the majority of our sample population only confirming, the Internet is undeniably being used increasingly as a preferred resource by the world around us.

### Discussion of Risk Factors Can Be Improved

Discussing risk factors with cancer patients is a sensitive topic that must be approached ethically by healthcare professionals. The survey concluded that 65% of the respondents stated they know and understand different risk factors for cancer. Additionally, only 53% of the respondents claimed they can communicate reliably with healthcare professionals regarding cancer prevention and care. Our study has found that more often than not the other 47% turn to the Internet for the information they found lacking, or felt uncomfortable with discussing cancer prevention and care with their provider.

### Patients Prefer In-Person to Online Appointments

With the rapid influx of online health-related resources, Telehealth is now a key player in the healthcare community. When the public was asked, “Would you prefer in-person consultations or telehealth for cancer care?” 64% responded in-person, while 18% responded via telehealth. Although most respondents still prefer in-person consultations or care, the public still refers to the Internet as a resource for health-related information. Another question led to 66% of the respondents agreeing that they know where to access cancer prevention information. Health-related information provided to patients over the Internet provides cancer patients with resources for their care and treatment.

### Residents in Urban, Rural, and Suburban Areas Share Somewhat Similar Preferences

Our one-way ANOVA, an analysis of variance to determine differences among means, revealed a significant effect of the differences between respondents’ geographic location and preferences on telehealth, F(3,181) = 2.85, p = 0.0387. Statistical significance is determined using a p-value threshold of 0.05, corresponding to a 95% level of confidence. The effect size, eta squared (η²), was 0.045, indicating a small effect. Our team conducted a generalized linear regression and found statistically significant and positive correlations when assessing whether those of different geographic regions use telehealth with their provider on a regular basis. We found a 177% increase in the likelihood of telehealth preference (p = 0.004) for urban residents, a 180% increase (p = 0.04) for suburban residents, and a 220% increase (p = 0.01) for rural residents. These results indicate that urban residents have the most statistically significant responses indicating that they use telehealth regularly. While, rural residents are second and suburban residents are last in terms of using telehealth regularly.

### Taking Steps to Reduce Cancer Risk Varies Based on Race

While spreading preventative information is important, actually taking these steps is what matters. A one-way ANOVA analysis revealed a significant effect of the differences between respondents’ self-identified racial background and whether they self-reported taking steps to reduce cancer risk, F(16,168) = 2.3, p = 0.0044, with a large effect size of η², = 0.179, showing a large effect size. The generalized linear regression found multiple statistically significant relationships between different racial backgrounds and the steps taken to reduce cancer risk. We found a positive relationship with a 263% increase in the likelihood of preventative steps taken (p = 2.9 E-06) for those of Asian descent. White Americans were found to have a 197% increase in the likelihood of preventative steps taken (p = 2.9 E-05). Black Americans reported a 175% increase (p = 0.0016). Lastly, Latina/o/x people were found to have a 135% increase (p = 0.028).

### Understanding of Cancer Prevention Methods Varies by Income Level

Income levels significantly influenced perceived understanding of cancer prevention methods, F(6, 178) = 2.93, p = 0.0093, with a medium effect size of η² =0.090. Generalized linear regression found statistically significant relationships between income levels and understanding cancer prevention methods. Respondents earning $0 - $25,000 showed a 158% increase in likelihood to understand cancer prevention methods (p = 0.02), $26,000 to $50,000 had a 188% increase (p = 0.004), $51,000 to $75,000 had a 168% increase (p = 0.011), $76,000 to $100,000 had a 216% increase (p = 0.0016), $100,000 to $200,000 had a 225% increase (p = 0.0008), and those earning over $200,000 had a 244% increase (p = 0.0006).

### Marital Status Impacts Ability to Communicate Reliably with Providers

A one-way ANOVA indicated a significant difference in communication reliability with providers based on marital status, F(5, 179) = 2.33, p = 0.044, with a medium effect size, eta squared (η²), of 0.061. Generalized linear regression showed that married respondents had a 198% increase in likelihood of reliable communication with providers (p = 0.002). Divorced respondents had a 144% increase (p = 0.048), single respondents had a 189% increase (p = 0.0035), and those in a domestic relationship had a 200% increase (p = 0.0055).

### Open-Ended Responses Correlate with Likert Scale Sentiments

The survey included written responses that allowed respondents to explain their actions and thoughts. The first qualitative question was: “Have you taken any steps in the last 12 months to reduce your risk of developing cancer?” Most participants stated that diet, sun protection, exercise, and regular screenings were the most common steps taken. However, for those who responded negatively, young age, lack of motivation, and busy schedules were the most common explanations for their uninterest in preventive measures. These demographic and cultural factors warrant further study in the future.

## Discussion

Our study results lend insight into how the general public receives, interprets, and acts upon healthcare information. Our research demonstrates the crucial role that information about preventive healthcare plays in the daily lives of adults in the US. It also reveals where there is room for improvement in levels of care and communication between patients and providers. Here we discuss these findings in relation to our research goals, examine their theoretical significance in health communication, and look at the practical implications for healthcare practitioners, policymakers, and digital health platforms.

### Public Perception of Online Health-related Information vs Provider Centered Care

Through the survey of the general public, our team was able to elucidate the public’s reported understanding and actions regarding cancer-related information. The fact that more respondents found it easier to obtain information online than from their doctors suggests a disconnect between information accessibility and perceived quality. Healthcare providers may bridge this gap by continuing to focus on the clarity and comprehensibility of the information they provide to patients (Ha & Longnecker, 2010).

With roughly 57 percent of respondents disagreeing that their doctor communicates with them about when cancer screenings should be conducted, it is not surprising that many people turn to online sources to find cancer information. Previous research suggests that physicians would not discuss cancer with patients due to various reasons, including inadequate time, physician forgetfulness, or a patient’s history of denial of care (Bhuyan et al., 2017). Considering this, open communication between physicians and patients is essential for a patient’s cancer knowledge and preparedness.

Cancer can be a highly emotional topic, thus the way it is spoken about in the media and in the clinical setting has the potential to impact treatment outcomes. A study conducted in 2010 concluded that emotions play a major role in influencing decision-making for cancer treatments or preventive care, even more than factual information (Zikmund-Fisher et al., 2010). Our survey found that only 53% of respondents claimed they can communicate reliably with healthcare professionals regarding cancer prevention and care. It may be the case that a lack of trust interferes with patients’ ability to communicate properly about cancer with their doctors.

One of the largest motivators for seeking out online health information is the patient’s needs not being adequately met by the healthcare system. The lack of access to consistent appointments and limited discussion time with their healthcare providers are just two factors that have led patients to turn to the Internet (Holmes, 2019). Regular screenings and discussions regarding future preventive behaviors are a vital part of a patient’s journey; thus, when left with no direction, patients turn to the Internet seeking the answers that they may not have received at their medical appointments. Given the room for improvement in patients’ health literacy and the fact that online information is not always accurate, there is an acute need for more open communication between doctors and their patients, as well as patient education on reliable Internet sources of medical information.

Ensuring healthcare information is accessible to all individuals is fundamental in promoting universal, equitable healthcare. Effective healthcare communication has the power to allow patients to be educated about their own health, facilitating a relationship of trust and ease between the patient and provider (Chichirez & Purcărea, 2018). Where there is trust and knowledge, mutual understanding and clarity are never far behind for both the patient and their provider.

### Communication of Cancer Risk Factors is Critical to Cancer Preventive Care

Direct interactions between patients and providers are vital to communicating different cancer risk factors. These risk factors are invaluable for patients to prevent and minimize cancer development. Common environmental stressors such as smoking, obesity, stress, and physical inactivity are rooted in up to 90% of all cancer diagnoses (Anand et al., 2008). The communication of the various risk factors plays a critical role in helping patients minimize their cancer development.

Previous research has shown that many people in the public have an understanding of the classic cancer risk factors (smoking, alcohol, diet) but do not correctly attribute lesser-known factors (environment, genetics, age) to cancer risk. Moreover, Ryan et al., 2015 found that many people falsely believe that cancer risk is not modifiable. While this can be true sometimes, certain behaviors can minimize the risk of cancer (Ryan et al., 2015). Our survey asked respondents whether they felt they understood cancer risk factors, with roughly 75% stating they agreed with that statement. While this number is a significant majority in the data, it does not give us concrete insight into whether or not they properly understand all risk factors. Response bias is a factor that should also be considered when examining these data.

Alternatively, previous literature has demonstrated that one’s perceived health status strongly impacts the frequency and diversity of searching for online health/cancer information (Xiao et al., 2014). Our findings suggest that many people are motivated to search for cancer information online, thus allowing them to implement preventive measures in their daily lives.

Increased outreach regarding the variety of risk factors for cancer and how to follow proper guidelines can be critical in minimizing cancer risk and progression of disease (White et al., 2014). By advocating for increased quantity and frequency of reliable resources regarding health-conscious behaviors online and encouraging providers, health organizations, and hospitals to recognize the importance of regularly communicating this information with their patients, the lowering of the mortality and incidence rate of most cancers can be expedited over the next several years (Jung et al., 2013).

### The Threat of Misinformation vs the Rise in Vital Online Prevention Information

As the accessibility of online cancer-related information has increased, more patients have been focusing on online sources to guide them throughout their care plans. Many informational websites regarding healthcare were originally intended for healthcare professionals but have now shifted to focus on preventive care and group support for patients (Benigeri & Pluye, 2003). Our results suggest people are using those resources and are taking steps to prevent cancer.

Although the public appears to be taking a step towards bettering their health and overall knowledge of cancer, with the help of the Internet, online medical information has the potential to have the opposite effect. The Internet contains information that is of high quality, allowing patients to make better health decisions. Still, it also offers inaccurate and often unreliable misinformation that can lead to increased levels of health-related anxiety (Robillard & Feng, 2017). The rise in misinformation campaigns and the spread of inaccurate health-related information threatens the newly symbiotic relationship between the Internet and the world of medicine.

The COVID-19 pandemic has additionally complicated patients’ ability to meet with doctors in person regarding preventive care (Haleem et al., 2021). This public health emergency led to the immediate adoption of telehealth for many healthcare visits. This increasing use of technology for health information has many implications for patients. While this option may provide fast and safe healthcare information, it may not fully meet patients’ needs and preferences. Increasing rates of public access to online cancer information can be correlated to increases in Internet usage over the last several decades. In the past, the communication of cancer-related information was restricted to receiving the information visually through informational pamphlets or verbally at a medical appointment. Now, telehealth appointments and online resources are rapidly on the rise, as shown by the continued influx of telemedicine visits and options available, even after the dust has settled from COVID-19 and in-person appointments are again available to most. Although COVID-19 may have initially triggered the rapid rise and development of telehealth appointments, some are now suggesting that telemedicine has proven to be sustainable, beyond the worldwide health crisis that made it what seemed to be a short-term necessity (Howie et al., 2022).

### Telehealth is Becoming a New Normal in Medical Care

Telehealth has many advantages that can improve patient health, such as improved efficiency and convenience, lower cost, and safety (Gajarawala & Pelkowski, 2021). Despite its benefits, telehealth also has costs that include difficulty maintaining the therapeutic relationship, poorer patient engagement, and a higher likelihood of missed diagnoses (Uscher-Pines et al., 2016). With the changing healthcare landscape, it is critical to understand the differing consequences of telehealth for patients and providers.

While telemedicine opportunities are continuously expanding, skepticism is still omnipresent, due in seemingly large part to the sensitive context of healthcare (Offermann et al., 2023). Telehealth, however, does have the potential to play a large role in the care plan for cancer patients. Not only do these appointments allow for alternative methods for receiving treatment and proper guidance from physicians, but they also provide flexibility for patients to attend appointments from the comfort of their homes. As cancer patients progress in their treatment plans, their overall health can begin to decline, thus requiring more appointments and travel time which makes it difficult to attend in-person appointments. Physicians who offer appropriate healthcare options may be able to reduce missed appointments (Snoswell & Comans, 2021). Whilst telehealth options are becoming more readily available, they are not replacements for in-person visits, as accessibility to the internet and devices to attend appointments may not be feasible for some patients (C. Y. Jiang et al., 2021).

Despite the rapid expansion of telehealth over the last several years, a substantial portion of the public (64%) still prefers in-person visits with their physician for their cancer care appointments. It is important to take into account patient preferences when offering telehealth to maintain a strong doctor-patient relationship. Shirke et al., 2020 have demonstrated that tele-oncology is becoming problematic for some cancer patients. Telehealth appointments also often require previous knowledge of technology, something many patients in older populations of different socioeconomic statuses, locations, and ages may struggle with. These results, coupled with previous literature, demonstrate the diverse range of factors that should be considered by the healthcare community while developing further telehealth options.

### Limitations

One of the limitations of this study is that the population of survey participants demographically tended to be on the younger side, with a mean age of 36, long before cancer and cancer screening is directly relevant to most of their lives. This necessitates the need for a future study targeting older demographics. This research study was also distributed as an online survey, meaning these respondents already have reliable Internet access and competency. A future analysis of online cancer preventive information should be administered to all people regardless of their access to the Internet and competency. This research serves as a pilot study to assess online cancer-preventive information in the post-COVID-19 world. Future research should also broaden the study to different cultures. We recommend future research to identify factors that motivate people to seek reliable online information.

### Avenues of Further Research & Next Steps in Literature

Effective cancer communication should be a priority for clinical and public health (T. C. Davis et al., 2002). To make it a priority in today’s society, social media and the Internet need to be utilized in a way where the resulting positives far outweigh the negatives. To learn to use social media to expand the reach of cancer-related prevention information and maximize the potential for a healthy lifestyle, organizations will need to formulate a strategic plan to ensure ongoing social media conversations rather than sporadic discussions spurred by events such as breast cancer awareness month. With further research, social media has the power to do amazing things for the oncology and healthcare community. Possible positive outcomes include raising awareness about clinical trials, promoting cancer prevention techniques, amplifying oncology information, pointing the public to reliable medical information, bringing diverse viewpoints into conversations, and educating medical colleagues regardless of geography (Morgan et al., 2022). Other possible next steps include pinpointing the susceptibility of different populations to low rates of health literacy and the spread of misinformation, as well as delving deeper into the most efficient and effective way to raise awareness for prevention strategies and other beneficial health-related behaviors.

## Conclusion

In conclusion, our findings shed light on healthcare information’s critical role in cancer-preventive behaviors. These insights provide a foundation for understanding how individuals seek, interpret, and utilize healthcare information. This research underscores the importance of clear and accessible healthcare information and the evolving landscape of healthcare communication. As we move forward, addressing these challenges and opportunities will be critical in promoting better overall patient health, outcomes, and well-being. Among the general public, many people prefer in-person appointments to telehealth. A majority of respondents in our survey indicated that they are not motivated by online cancer preventive information to take steps to limit their cancer risk. Additionally, patients acknowledged that they are often turning to online cancer information because their needs are not fully met by their providers. This analysis reveals that cancer information from one’s healthcare provider can coexist with online cancer information. Still, a patient should consult with their provider for the most accurate and personalized care.

## Data Availability

All data produced in the present study are available upon reasonable request to the authors

## Acknowledgements

We would like to acknowledge the Arizona Cancer Evolution Center and the Arizona Cancer Evolution Center (ACE) Scholars program at Arizona State University. The team’s support, mentorship, and guidance have been invaluable to this research process. We would also like to thank patient advocates Mindy Miller and Erin McGee Ferrell for so generously providing us with a patient’s perspective. We are grateful to Pamela Winfrey for inspiring conversations about communication and art in cancer. Our team is appreciative of the survey participants for allowing us to gauge the public’s opinions about this important topic.

